# Multi-level modeling of early COVID-19 epidemic dynamics in French regions and estimation of the lockdown impact on infection rate

**DOI:** 10.1101/2020.04.21.20073536

**Authors:** Mélanie Prague, Linda Wittkop, Annabelle Collin, Dan Dutartre, Quentin Clairon, Philippe Moireau, Rodolphe Thiébaut, Boris P. Hejblum

## Abstract

We developed a multi-level model of the French COVID-19 epidemic at the regional level. We rely on a global extended Susceptible-Exposed-Infectious-Recovered (SEIR) mechanistic model as a simplified representation of the average epidemic process, with the addition of region specific random effects. Combining several French public datasets on the early dynamics of the epidemic, we estimate region-specific key parameters conditionally on this mechanistic model through Stochastic Approximation Expectation Maximization (SAEM) optimization using `Monolix` software. We thus estimate the basic reproductive numbers by region before lockdown (with a national average at 2.81 with 95% Confidence Interval [2.58; 3.07]), attack rates (i.e. percentages of infected people) over time per region which range between 1.9% and 9.9% as of May 11^*th*^, 2020, and the impact of nationwide lockdown on the infection rate which decreased the transmission rate by 76% towards reproductive numbers ranging from 0.63 to 0.73 at the end of lockdown across regions. These results confirm the low population immunity, the strong effect of the lockdown on the dynamics of the epidemics and the need for further intervention when lifting the lockdown to avoid an epidemic rebound.

## 1 Introduction

In December 2019, grouped pneumonia cases have been described in the Hubei province, China and SARS-CoV-2 was identified on January, 7^th^ as the cause of this outbreak (Li et al., 2020; Zhu et al., 2020). SARS-CoV-2 causes the viral disease which has been named COVID-19 (World Health Organization, 2020b). SARS-CoV-2 rapidly spread all over the world and the pandemic stage was declared on March 11^th^ by the World Health Organization (2020c). On May 11^th^, over 4,006,257 cases (in accordance with the applied case definitions and testing strategies in the affected countries) including 278,892 deaths had been reported (World Health Organization, 2020a). In France, the first case was declared on January, 24^th^ (Bernard-Stoecklin et al., 2020), nationwide lockdown was initiated on March 17^th^ until May 11^th^. On that date of lockdown lift, *Santé Publique France* reported 139,519 confirmed cases and 26,643 deaths due to COVID-19 (among which 16,820 occurred at the hospital) in France.

COVID-19 includes non-specific symptoms such as fever, cough, headache, and specific symptoms such as alteration of smell and taste (Gane et al., 2020; Lechien et al., 2020; Tong et al., 2020). The virus is transmitted through droplets and close unprotected contact with infected cases. The majority (around 80 %) of infected cases have a mild form (upper respiratory infection symptoms) without specific needs in terms of care. Around 20 % of cases need hospitalization and among those are severe forms (severe respiratory distress) which will need to be admitted to intensive care units (ICU) with potential need of mechanical ventilation. The percentage of patients in need for ICU care varies between 5 % reported from China (Guan et al., 2020) and 16 % reported from Italy (Grasselli et al., 2020). The number of ICU beds in France was 5,058 at the end of 2018 (DREES, 2019). Thus, the availability of ICU beds with mechanical ventilation is one of the major issues as facilities were not prepared to deal with the potential increase of the number of patients due to this pandemic.

Unprecedented public-health interventions have been taken all over the world (Kraemer et al., 2020) to tackle this epidemic. In France, interventions such as heightening surveillance with rapid identification of cases, isolation, contact tracing, and follow-up of potential contacts were initially implemented. But as the epidemic continued growing, comprehensive physical distancing measures have been applied since March 15^th^, 2020 including closing of restaurants, non-vital business, schools and universities *etc*, quickly followed by state-wide lockdown on March 17^th^ 2020. The French president has announced on April 13^th^ 2020, a progressive lifting of the lockdown from May 11^th^ 2020 onwards. In Wuhan (Hubei, China), the extremely comprehensive physical distancing measures in place since January 23^rd^ have started to be relaxed after 2 months of quarantine and lifted completely on April 8^th^ 2020 (Tian et al., 2020; Wu and McGoogan, 2020). Interestingly, these interventions have been informed by mathematical models used to estimate the epidemic key parameters as well as unmeasured compartments such as the number of infected people.

Several approaches have already been proposed to model and forecast the COVID-19 epidemic using compartment models (Fang et al., 2020; Tang et al., 2020; Wang et al., 2020), or agent based models (Di Domenico et al., 2020; Ferguson et al., 2020; Wilder et al., 2020), its potential impact on intensive care systems (Fox et al., 2020; Massonnaud et al., 2020), and to estimate the effect of containment measurements on the dynamics of the epidemic (Magal and Webb, 2020; Prem et al., 2020). Most of those rely on simulations with fixed parameters and do not perform direct statistical estimations from incident data (Massonnaud et al., 2020; Roux et al., 2020). Roques et al. (2020) used French national data but did not use a multi-level approach to model the epidemic at a finer geographical granularity. Yet, the dynamics of the epidemic can be very heterogeneous between regions inside a given country resulting in tremendous differences in terms of needs for hospital and ICU beds (Massonnaud et al., 2020; Roux et al., 2020). Moreover, the data collection yields noisy observations, a problem that we deal with thanks to statistical modeling of the observation process rather than directly altering the data by e.g. smoothing such as in Roques et al. (2020). Salje et al. (2020) also use several models and statistical analyses to estimate the dynamics of the epidemics in France.

Here, we use public data from the COVID-19 outbreak in France to estimate both the dynamics of the COVID-19 epidemic at the French regional level and the impact of the nationwide lockdown. We propose a global model including all potential individual states associated with a multi-level approach where each region is considered as a statistical unit. A key feature of our modeling strategy is to clearly separate three layers in our model: i) the observation model that will account for the data collection process and measurement error, ii) the mechanistic model which describes the average disease process and its evolution through a system of dependent Ordinary Differential Equations, and iii) the statistical estimation model which leverages the flexibility of random effects to accommodate the heterogeneity between French regions and estimate both national and region-wise quantities of interest (see Prague et al. (2012)). By pooling information across regions, we augment the data to increase estimate precision, while random effects explicitly acknowledge variations between regions. Their heterogeneity is partly due to different initial epidemic states, such as differences in virus introduction timelines or the occurrence of super-spreading events (Roux et al., 2020), but it also encompasses different intrinsic characteristics such as population density, age structure, and transportation habits that influence disease spread through each region-specific population.

The mechanistic model is a SEIRAH model, which is an extended Susceptible - Exposed - Infectious - Recovered (*SEIR*) model accounting for time-varying population movements, non-reported infectious subjects (*A* for unascertained) and hospitalized subjects (*H*), as proposed by Wang et al. (2020) to originally model the COVID-19 epidemic in Wuhan. Parameters from this model are estimated at the regional scale using a multi-level approach which borrows information across regions, increasing the amount of data and thereby strengthening the inference while allowing for local disparities in the epidemic dynamics. In addition, we use forward simulations to predict the effect of non-pharmaceutical interventions (NPIs) (such as lift of lockdown) on the evolution of the epidemic. Section 2 introduces the data, the model and the necessary statistical tools, Section 3 presents our results, and Section 4 discusses our findings and their limits.

## 2 Methods

Because epidemics spread through direct contacts, their dynamics have a strong spatial component. While traditional compartment models do not account for spatiality, we propose to take it into account by: i) modeling the epidemic at a finer, more homogeneous geographical scale (this is particularly important once lockdown is in place); ii) by using a multi-level approach with random effects across French regions which allows each region to have relatively different dynamics while taking all information into account for the estimation of model parameters iii) aligning the initial starting time of the epidemic for all regions. The starting date in each region was defined as the first date with incident confirmed cases of COVID-19 directly followed by 3 additional consecutive days with incident confirmed cases as well. This criterion of 4 consecutive days with incident cases is needed in particular for the *Île-de-France* region which had 3 consecutive days with 1 imported confirmed case in late January which did not lead to a spreading outbreak at that time.

### 2.1 Data sources

Open-data regarding the French COVID-19 epidemic is currently scarce, as the epidemic is still unfolding. *Santé Publique France* (SPF) in coordination with the French regional health agencies (*Agences Régionales de Santé* – ARS) has been reporting a number of aggregated statistics at various geographical resolutions since the beginning of the epidemic. During the first weeks of the epidemic in France, SPF was reporting the cumulative number of confirmed COVID-19 cases with a positive PCR test. Other French surveillance resources such as the *Réseau Sentinelles* (Valleron et al., 1986) or the SI-VIC (*Système d’information pour le suivi des victimes*, maintained by the ANS – *Agence du Numérique en Santé*, formerly named ASIP) database quickly shifted their focus towards COVID-19, leveraging existing tools to monitor the ongoing epidemic in real time, making as much data available as possible (whith respect to data privacy regulation). In this study, we combined data from three different data sources: i) the daily release from SPF, from which we computed the daily incident number of confirmed COVID-19 cases (i.e. with a positive PCR test) in each region; ii) the SI-VIC database that started recording COVID-19 patients hospitalizations in French public and private hospitals on March, 13^th^, from which we computed the daily incident number of COVID-19 hospitalization in each region. All cases are either biologically confirmed or present with a computed tomographic image highly suggestive of SARS-CoV-2 infection; iii) the *Réseau Sentinelles* which started estimating the weekly incidence of COVID-19 in each French region on March 16^th^. We used the *Réseau Sentinelles* network’s weekly incidence estimates of symptomatic cases (including non confirmed cases) to set the ratio between ascertained and unascertained cases in each region (later denoted as *r*). Table 1 presents these observed data. Of note, we studied the epidemic in the 12 Metropolitan French regions – excluding the Corsican region (*Corse*) which exhibits different epidemic dynamics, possibly due to its insular nature.

**Table 1:** Description of the COVID-19 epidemic by French region (on available data). The epidemic start date is defined as the first day followed by three consecutive days with at least 1 confirmed case. National totals are displayed for France, except for *r*_*s*_ which is a weighted average.

| Geographical region | Epidemics start date | Observed cumulative Ascertained cases until 2020-03-25 | Observed cumulative Hospitalized cases until 2020-05-11 | Population on 2018-12-31 | Percentage of ascertained cases from *Réseau Sentinelles* $r_s$ |
| --- | --- | --- | --- | --- | --- |
| *Auvergne-Rhône-Alpes* | 2020-03-02 | 1,846 | 9,072 | 8,032,377 | 0.03 |
| *Bourgogne-Franche-Comté* | 2020-03-02 | 1,506 | 4,520 | 2,783,039 | 0.07 |
| *Bretagne* | 2020-03-02 | 525 | 1,528 | 3,340,379 | 0.02 |
| *Centre-Val de Loire* | 2020-03-05 | 450 | 2,777 | 2,559,073 | 0.03 |
| *Grand Est* | 2020-03-02 | 4,921 | 15,381 | 5,511,747 | 0.05 |
| *Hauts-de-France* | 2020-03-02 | 1,523 | 7,770 | 5,962,662 | 0.02 |
| *Île-de-France* | 2020-03-02 | 6,789 | 36,367 | 12,278,210 | 0.04 |
| *Normandie* | 2020-03-10 | 575 | 2,053 | 3,303,500 | 0.02 |
| *Nouvelle-Aquitaine* | 2020-03-04 | 785 | 2,451 | 5,999,982 | 0.03 |
| *Occitanie* | 2020-03-02 | 899 | 3,171 | 5,924,858 | 0.03 |
| *Pays de la Loire* | 2020-03-04 | 338 | 2,366 | 3,801,797 | 0.01 |
| *Provence-Alpes-Côte d’Azur* | 2020-03-02 | 1,595 | 6,127 | 5,055,651 | 0.05 |
| *France* | NA | 21,752 | 93,583 | 64,553,275 | 0.034 [0.01; 0.07] |

### 2.2 Model

#### 2.2.1 Structural model of the epidemic

Wang et al. (2020) extended the classic SEIR model to differentiate between different statuses for infected individuals: ascertained cases, unascertained cases and cases quarantined by hospitalization. The model, assuming no population movement, is presented in Figure 1. The population is divided into 6 compartments: susceptible *S*, latent *E*, ascertained infectious *I*, unascertained infectious *A*, hospitalized *H*, and removed *R* (i.e. both recovered and deceased). This model assumes that infections are well-mixed throughout the population, ignoring any spatial structure or compartmentalization by population descriptors such as age for instance. Such assumptions make it particularly relevant to infer the dynamics of the French epidemic at the regional level (a finer geographical scale at which such hypotheses are more likely to hold). Figure 1 illustrates the dynamics between those 6 compartments that are characterized by the following system of six Ordinary Differential Equations (ODE):

**Figure 1:**
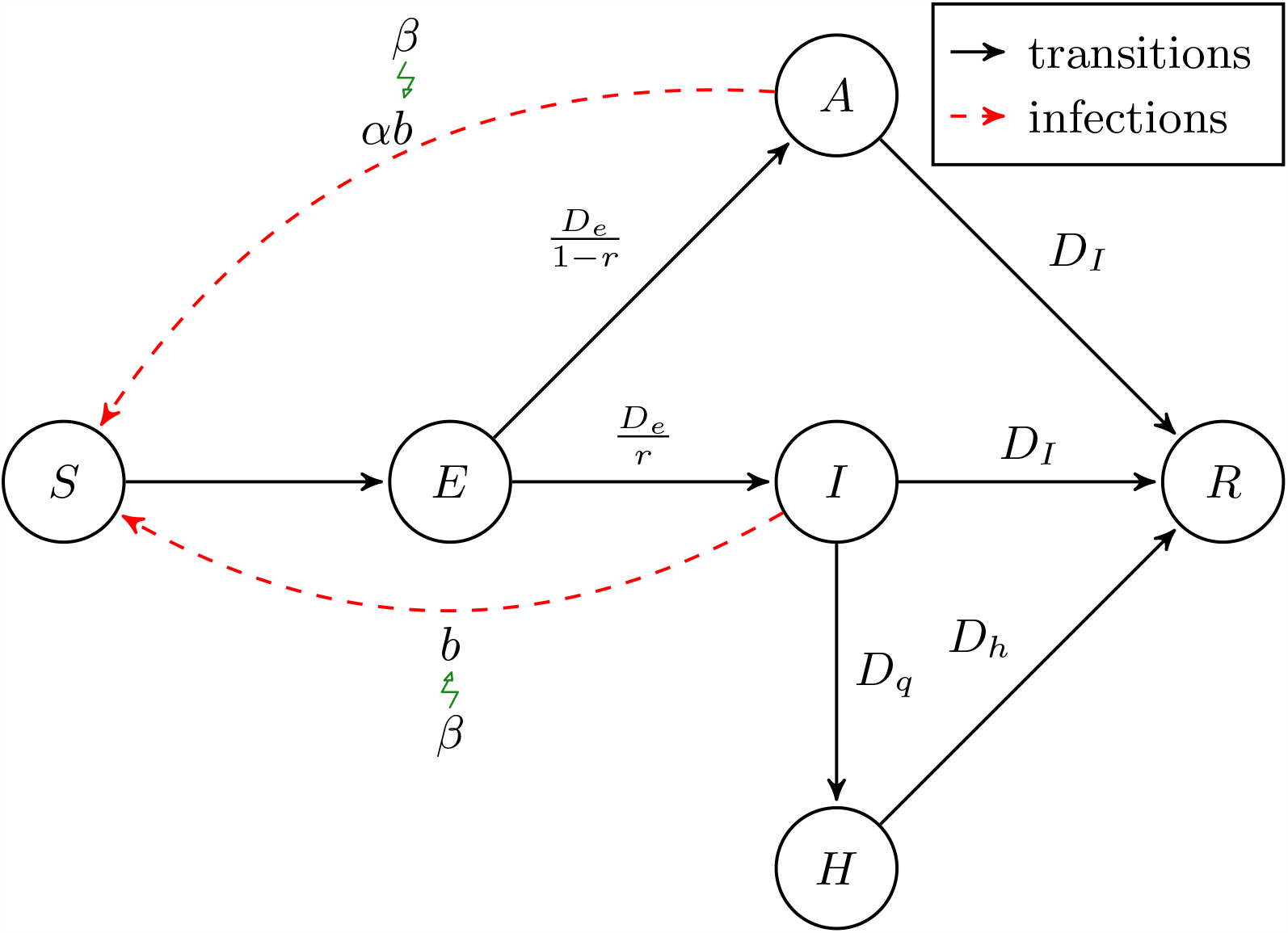
SEIRAH model representation – adapted from Wang et al. (2020)

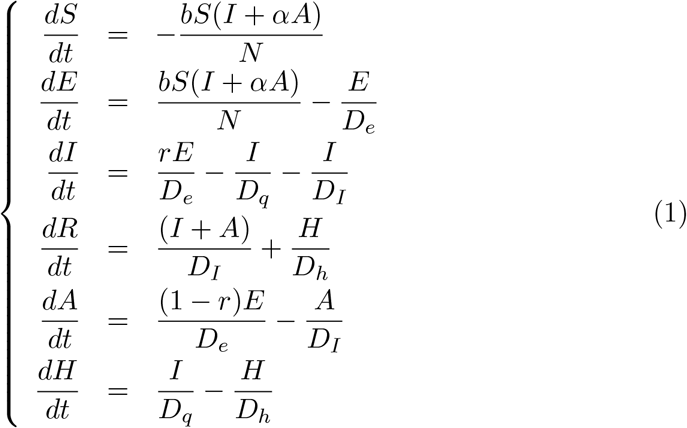

Model parameters are described in Table 2. Of note, given a combination of parameters and initial states of the system *ξ* = (*b, D*_*q*_, *r, α, D*_*e*_, *D*_*I*_, *D*_*h*_, *N, S*(*t* = 0), *E*(*t* = 0), *I*(*t* = 0), *R*(*t* = 0), *A*(*t* = 0), *H*(*t* = 0)), using a solver of differential equations, it is possible to deterministically compute at any time *t* the quantities *S*(*t, ξ*), *E*(*t, ξ*), *I*(*t, ξ*), *R*(*t, ξ*), *A*(*t, ξ*), and *H*(*t, ξ*).

**Table 2:** Parameters of the SEIRAH model.

| Parameter | Interpretation | Value | References |
| --- | --- | --- | --- |
| $b$ | Transmission rate of ascertained cases | region specific | *estimated* |
| $r$ | Ascertainment rate | region specific | *Réseau Sentinelles* |
| $\alpha$ | Ratio of transmission between $A$ and $I$ | 0.55 | Li et al. (2020) |
| $D_e$ | Latent (incubation) period (days) | 5.1 | Lauer et al. (2020) |
| $D_I$ | Infectious period (days) | 2.3 | Li et al. (2020); Wang et al. (2020) |
| $D_q$ | Duration from $I$ onset to $H$ (days) | region specific | *estimated* |
| $D_h$ | Hospitalization period (days) | 30 | Li et al. (2020); Wang et al. (2020) |
| $N$ | Population size | region specific | INED (2020) |
| $\beta$ | Lockdown effect | - | *estimated* |

#### 2.2.2 Observation model

##### Observation processes

In our case, none of the compartments of the system are directly observed: the only observations considered are i) the number of daily incident infectious ascertained cases denoted *Y*^1^, and ii) the number of daily incident hospitalized infectious cases denoted *Y*^2^. These observations are available both before and after the initiation of lockdown. Those two quantities are modeled in Equation (1) respectively as observations from the 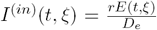 and 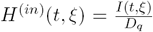 random variables, which are the numbers of new incident cases at time *t* given the parameters *ξ* in compartment *I* and *H* respectively. Because these are count processes, we propose to model their observations *Y*^1^ and *Y*^2^ with Poisson likelihoods:

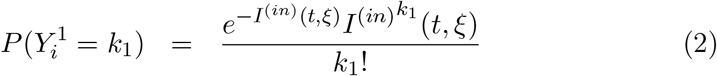

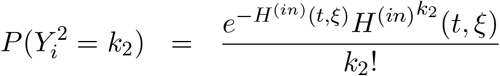

where *k*_1_ and *k*_2_ are the respective numbers of cases.

##### Initial values

The initial states of all compartments at the date of epidemic start (*t* = 0) for region *i* are also important drivers of the region dynamics. Some of them can be directly approximated such as *R*_0*i*_ = *R*_*i*_(*t* = 0) = 0, while others are supposedly observed such as *H*_0*i*_ and *I*_0*i*_. However, because data collection has been somewhat inconsistent between regions (e.g. with some regions being late in reporting their first cases), it is probably not adequate to directly initialise using these observations (for example *Normandie* has a higher *I*_0*i*_ than *Grand-Est* while the epidemic was much larger in *Grand-Est*). Finally, others compartments (namely *A* and *E*) are not directly observed. We used a two-step strategy based on a Kalman-filter estimation approach to find a credible initial value for each initial states (*I*_0*i*_, 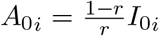, and *H*_0*i*_) at time t=0 (see Section S2.1 of Web Appendix 1). Moreover, *E*_0*i*_ = *E*_*i*_(*t* = 0) are estimated as model parameters (see Section 2.2.3). Finally, because the total population size is equal to *N*_*i*_, we have *S*_0*i*_ = *S*_*i*_(*t* = 0) = *N*_*i*_ − *E*_0*i*_ − *I*_0*i*_ − *R*_0*i*_ − *A*_0*i*_ − *H*_0*i*_.

#### 2.2.3 Statistical multi-level model

The goal of this study is to model the epidemic of COVID-19 in France, but at the regional level using a multi-level approach. This is done using a mixed effect model. In this inference framework, baseline parameters governing the dynamics of the epidemic in each region are assumed to be drawn from a shared distribution which allows for heterogeneity between regions, known as the random effects. We use the log-normal distribution for all parameters to ensure their positivity during estimation. Because public health policies changed over the time period of observation of the epidemic, we incorporate explanatory covariates such as physical distancing by lockdown. In order to substantiate the parametric shape chosen for modeling the disease transmission *b*, we have investigated the use of a Kalman-based estimation strategy (Bensoussan, 2018; Simon, 2006) applied to the dynamic system of eq. (1). The full strategy is presented in Section S2.1 of Web Appendix 1 and its results (detailed in Section S2.2.2 of Web Appendix 1) validate a choice of a step function with four breaks as described below. This choice was further justified by a model selection procedure minimizing the AIC. Thus, denoting *τ*_*i*_ the lockdown time in region *i*, we have ∀*i* = 1, …, 12 (where *i* is the region identifier):

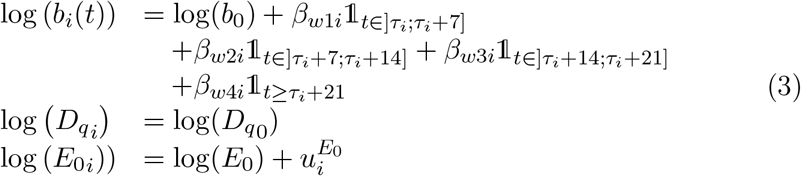

With 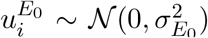, and 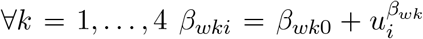 and 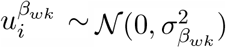. The parameters (*b*_0_, 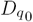, *E*_0_, *β*_*w*10_, *β*_*w*20_, *β*_*w*30_, *β*_*w*40_) are mode shared values across regions. Of note, this is different from the French mean value which is obtained by a weighted mean of the population parameters depending on the size of each region. The inter-region random-effect 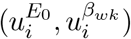 are normally distributed and assumed independent. So the vector of parameters in the model for each region is *ξ*_*i*_=(*b*_*i*_, *D*_*q*_*i, β*_*w*1*i*_, *β*_*w*2*i*_, *β*_*w*3*i*_, *β*_*w*4*i*_, *r, α, D*_*e*_, *D*_*I*_, *D*_*h*_, *N*_*i*_, *S*_0*i*_, *E*_0*i*_, *I*_0*i*_, *R*_0*i*_, *A*_0*i*_, *H*_0*i*_). *β*_*wk*0_ can be interpreted through *K*_*k*_ = 1− exp(− *β*_*wk*0_) which is the percentage by which transmission is decreased after the start of lockdown during the *k*^th^ week (or from the fourth week to the end of lockdown for k=4). The coefficients *β* = (*β*_*w*10_, *β*_*w*20_, *β*_*w*30_, *β*_*w*40_) are expected to be negative as lockdown aims at reducing transmission. Interestingly, with our approach, we can evaluate whether or not there is a statistically significant effect of lockdown on the transmission by testing *β*_*wk*0_ = 0 using a Wald test.

### 2.3 Inference

#### 2.3.1 Estimation

##### Region-specific model parameters

Based on the results from the theoretical identifiability analysis of the structural model of the epidemic from Equation (1) (see Section S1 of Web Appendix 1), we estimate the parameters (*b*_*i*_,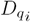) as well as the initial state (*E*_0*i*_) and intervention effects (*β*_*w*1*i*_, *β*_*w*2*i*_, *β*_*w*3*i*_, ans *β*_*w*4*i*_) when the epidemic begins being reported in each region *i*. We used the Monolix software version 2019R2 (Lixoft SAS, 2019) to estimate those seven parameters by maximizing the likelihood of the data given the model and the other fixed parameters. This software relies on a frequentist version of the Stochastic Approximation Expectation Maximization (SAEM) algorithm (Delyon et al., 1999) and standard errors are calculated via estimation of the Fisher Information Matrix (Kuhn and Lavielle, 2005), which is derived from the second derivative of the log-likelihood evaluated by importance sampling. In addition, we use profile-likelihood to confirm that no further information can be gained from the data at hand on parameters *α, D*_*e*_ and *D*_*I*_ by running the SAEM algorithm multiple times while setting these parameters to different values and obtaining similar maximum likelihood values (meaning more data would be needed to be able to estimate those parameters). During inference, practical identifiability of the model is evaluated by the ratio of the minimum and maximum eigenvalues of the Fisher Information Matrix, that will be referred as “convergence ratio” in the reminder of the manuscript. Convergence of the SAEM algorithm was assessed by running multiple SAEM chains and checking that they all mix around similar probability distributions.

##### Compartments dynamics

We are particularly interested in the trajectories of the model compartments. We use Monte Carlo methods (parametric bootstrapping) to compute the confidence intervals accounting for the uncertainty in estimating the structural and statistical model parameters. For all compartments *C*(*t, ξ*_*i*_) (*C* being *S, E, I, R, A*, or *H*) the 95% confidence interval is estimated by sampling from the posterior distributions of the model parameters to simulate 1,000 trajectories, and taking the 2.5% and 97.5% percentiles of these simulated trajectories. We also added to it the error measurement given by the Poisson distribution of the outcomes. Another outcome of interest is the number of death (D). This quantity is not specifically modeled by our mathematical structural model. However, it is possible to roughly approximate it by assuming that it represents a percentage of the removed cases *R*(*t, ξ*_*i*_). Based on the estimation of the Infection Fatality Ratio (IFR) from Roques et al. (2020), we get a rough estimation of *D*(*t, ξ*_*i*_) as 0.5% of *R*(*t, ξ*_*i*_). ‘

##### Effective reproductive number

For each region, we compute the effective reproductive number *R*_*e*_(*t, ξ*_*i*_) as a function of model parameters:

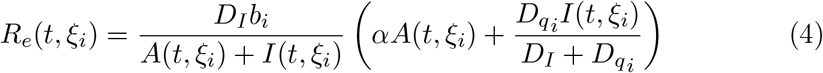

When individuals are homogeneous and mix uniformly, *R*_*e*_(*t, ξ*_*i*_) is defined as the mean number of infections generated during the infectious period of a single infectious case in the region *i* at time *t*. This is the key parameter targeted by NPIs. We compute analytically its 95% confidence interval by accounting for 95% confidence interval of all compartments.

##### Asymptomatic proportion

At a given time *t* the number of incident unascertained cases is equal to the sum of two populations, the number of incident non-tested symptomatic individuals (NT) and the number of incident non-tested asymptomatic individuals (AS):

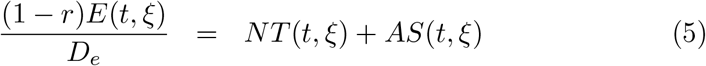

where *r* is the proportion of cases tested positive. Collection of data from general practitioners through the re-purposing of the *Réseau Sentinelles* network to monitor COVID-19 provides a weekly estimation of the number of incident symptomatic cases (tested or not tested) that we previously called *r*_*s*_. This quantity is given over a week but can be evaluated daily by averaging:

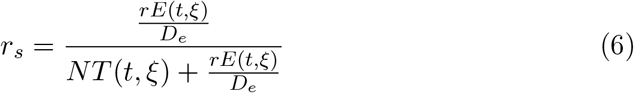

where *r*_*s*_ represents the proportion of infected cases seeing a general practitioner. Combining equations (5) and (6) allows to compute the incident number of asymptomatic cases as a function of the compartment *E*:

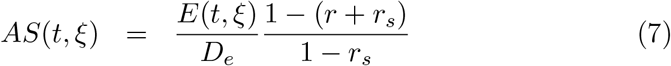

##### Short-term predictions of attack rates

We predict the proportion of infected individuals among the population in each region at a given date by computing: [*E*(*t, ξ*_*i*_) + *A*(*t, ξ*_*i*_) +*I*(*t, ξ*_*i*_) + *H*(*t, ξ*_*i*_) + *R*(*t, ξ*_*i*_)] */N*_*i*_ (neglecting the deaths).

## 3 Results

### 3.1 Estimation of the regional epidemic dynamics

#### Data fitting

On May 11^th^, the cumulative number of ascertained cases was 21,752 (with available data until March 23^rd^ only) and the cumulative number of hospitalized cases was 93,583, see Table 1 for a regional breakdown. Our SEIRAH model fits the data well as can be seen in Figure 2. Moreover, the stability of the estimates is good with a convergence ratio of 1.7 (see Section 2.3.1), corroborating the good identifiability of the estimated parameters (see Section S1 of Web Appendix 1). Table 3 provides the regional estimates of the baseline number of exposed cases in each region *E*_0*i*_. Of note, it is significantly higher in regions in which the epidemics took off, such as *Île-de-France* and *Grand Est*. The transmission rate *b*_*i*_ and the time from illness confirmation to hospitalization 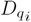 is estimated to be the same in each region and equal to 2.23 (sd. 0.003) and 0.36 (sd. 0.01). The parameter 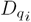 is lower than 1 because of the data collection procedure. Especially in the beginning of the epidemics, there was often a delay between COVID-19 confirmation and hospitalization. Actually, a consequent part of the patients were probably tested after being hospitalized.

**Figure 2:**
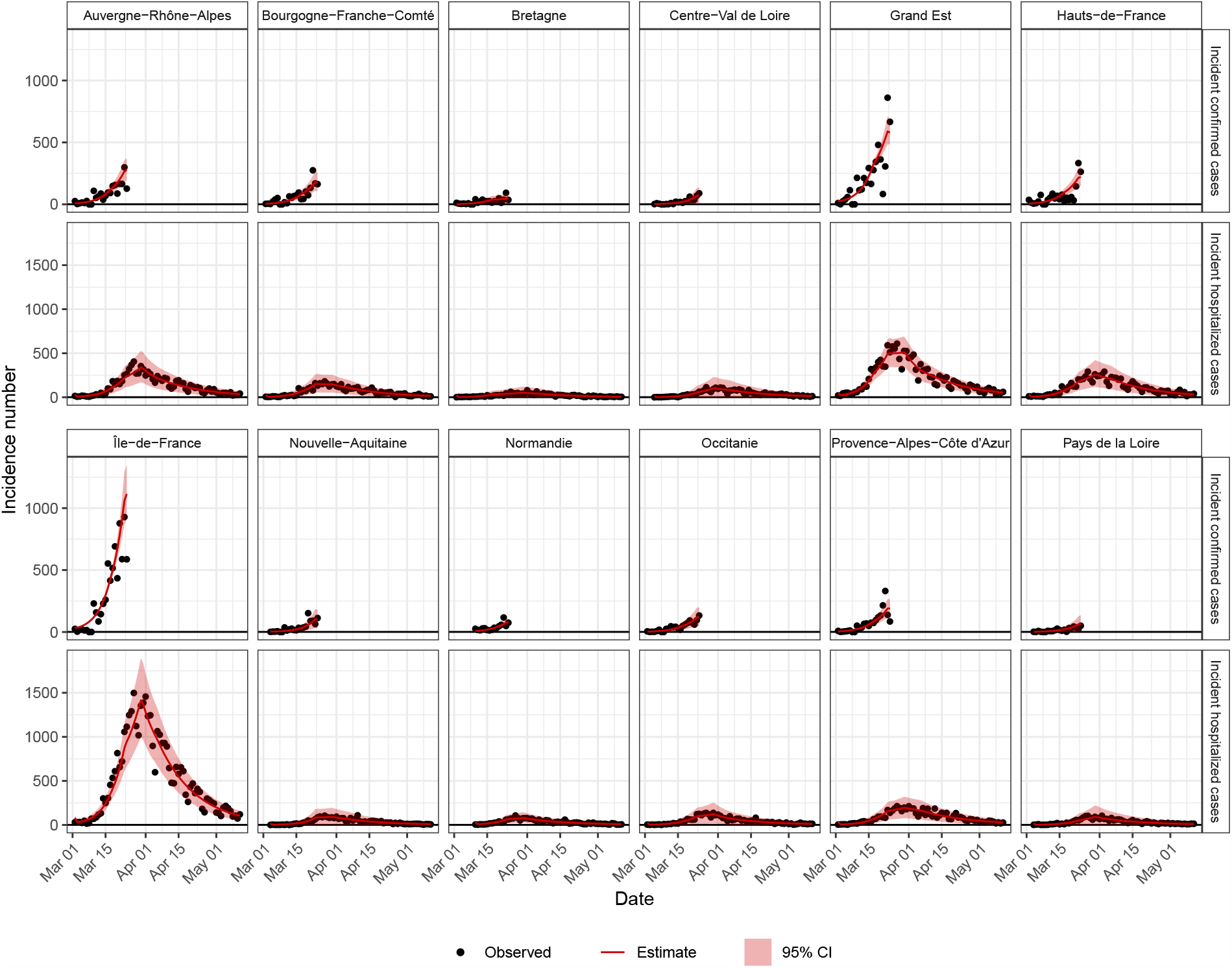
Fitting curves of *I*^(*in*)^ and *H*^(*in*)^ with the SEIRAH model.

**Table 3:**
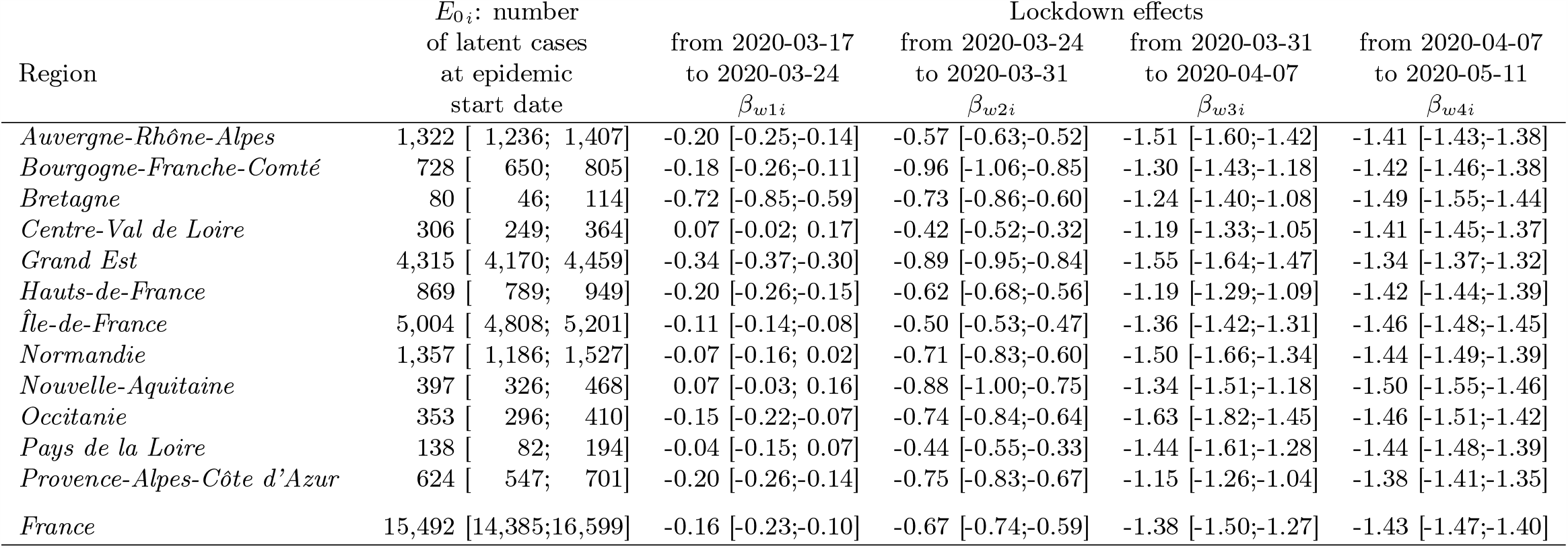
Estimation of region-wise model parameters and national weighted averages.

#### Prediction abilities

To evaluate the validity of our structural ODE model (1) and of our inference results, we compare the aggregated predictions of the number of both incident ascertained cases and incident hospitalized cases at the national French level to the daily observed incidences (that are still publicly available from SPF at the country level, even after March 25^th^ – while incident ascertained cases are not openly available at the regional level after March 25^th^). We also derive the number of cumulated deaths from our model and compare it to the reported number of deaths by SPF. Figure 3 displays both predictions and observations. All data series display a good agreement although the model was not fitted using the same exact sources of data and the observations used were only partially containing these data -i.e. no confirmed cases after March 25^th^ and no data on death.

**Figure 3:**
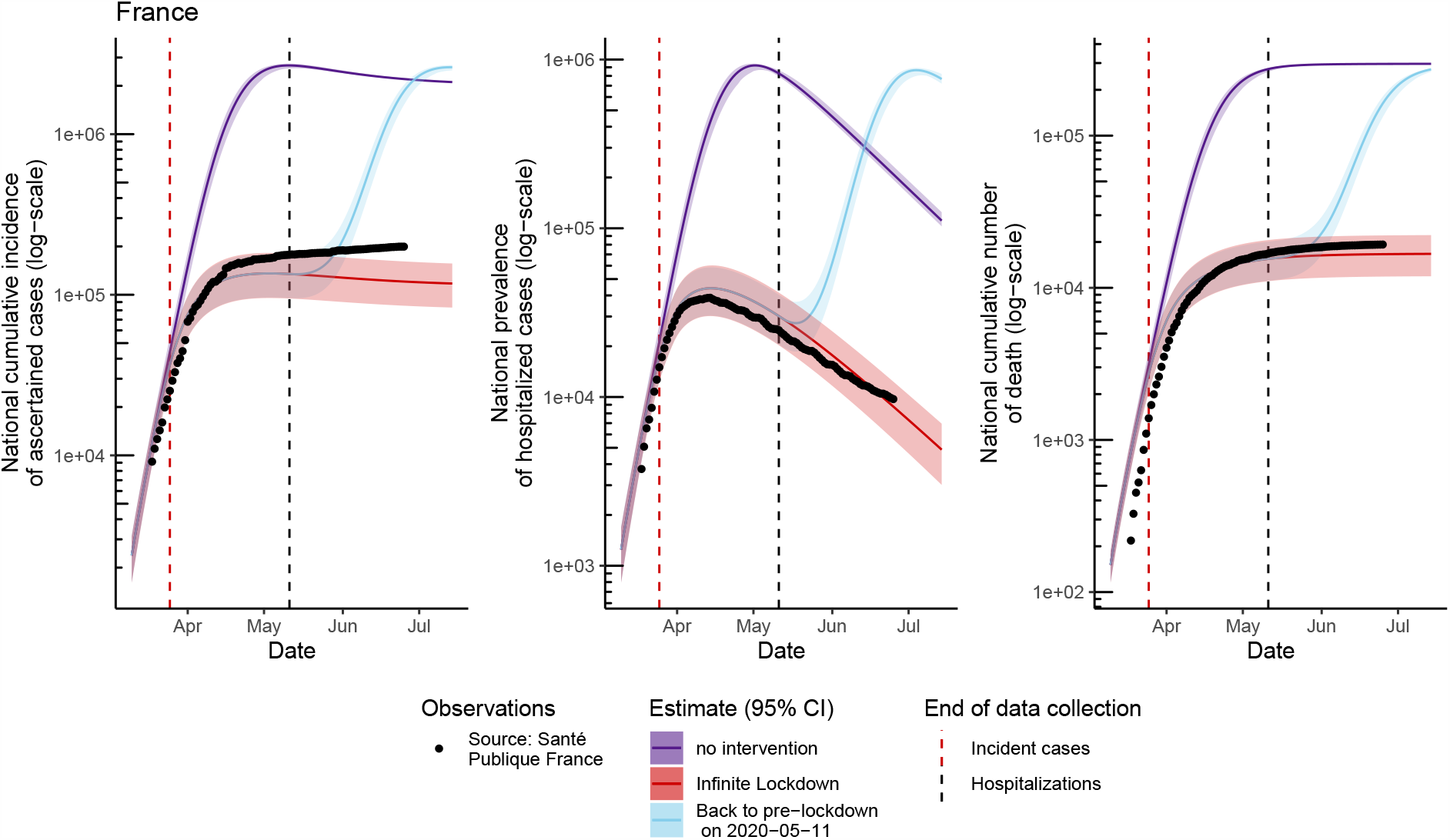
Observed incident number of ascertained cases, hospitalizations and deaths in France compared to fitted/predicted ones with the model, based on estimations with data collected up to March 25^th^ for incident number of ascertained cases and hospitalizations up to May 11^th^.

#### Evolution of the epidemic without intervention

It is also interesting to predict the percentage of infected individuals in each region at future dates, corresponding to attack rates. In the absence of any NPI, we predict that 92.1% [91.9%;92.3%] of the total French population would end up being infected by the SARS-CoV-2 by the end of the epidemic. In this scenario, we predict that the peak of the epidemic would have been around April 19^th^ 2020 and that the epidemic would have finally gone extinct around June 28^th^ 2021. At the peak of hospitalization, about 922,786 [844,634; 943,681] individuals would have required simultaneous hospitalization for COVID-19. Also, under this model assumptions, we crudely estimate that the number of death prevented by the implemented NPIs over the period of lockdown is 258,573 [239,112; 266,960].

#### Proportion of asymptomatic cases

With the current model, we have *D*_*e*_ = 5.1 and *r* = *r*_*s*_ = 3.3% [1.2%; 6.6%]. Thus we estimate that the percentage of asymptomatic infectious individuals is : 18.1% [16.7%; 19.2%]. This estimate is in line with estimation obtained from active surveillance data on the Diamond Princess cruise ship (Mizumoto et al., 2020).

### 3.2 Estimation of the lockdown effect

#### Change of transmission rate during lockdown

The parameters *β*_*wk*_ for *k* = 1 …, 4 measure the effect of the lockdown. They are all significantly smaller than 0 (p< 0.001) such that the lockdown reduced the transmission rate of COVID-19 from the first week onward. The transmission is decreased by 14.8%[9.5%; 20.5%] the first week, by 48.8%[44.6%; 52.3%] the second week, by 74.8%[71.9%; 77.7%] the third week, and by 76.1%[75.3%; 77.0%] thereafter. A breakdown by region is available in Table 3.

#### Effective reproductive number

The above quantities directly impact the effective reproductive number as described in Table 4. The overall effective reproductive number for France is 2.81 [2.58;3.07] before the lockdown, 2.42 [2.01;2.90] during the first week of lockdown, 1.46 [1.24;1.72] during the second week of lockdown, 0.71 [0.59;0.87] during the third week of lockdown and 0.67 [0.58;0.78] thereafter. Figure 4 displays the effective reproductive number trajectories in each region.

**Table 4:** Estimation of the effective reproductive ratios *R*_*e*_ during each of the 3 considered periods (before lockdown, during the first week of lockdown, and beyond 1 week of lockdown) for each region with 95% confidence intervals and national weighted averages.

| Geographical region | $R_e$<br>(before<br>confinement)<br>on<br>2020-03-15 | $R_e$<br>(during<br>confinement)<br>first week<br>on 2020-03-22 | $R_e$<br>(during<br>confinement)<br>second week<br>on 2020-03-29 | $R_e$<br>(during<br>confinement)<br>third week<br>on 2020-04-05 | $R_e$<br>(during<br>confinement)<br>after<br>on 2020-05-11 |
| --- | --- | --- | --- | --- | --- |
| *Auvergne-Rhône-Alpes* | 2.81 [2.62;3.01] | 2.31 [1.96;2.73] | 1.59 [1.38;1.84] | 0.62 [0.53;0.74] | 0.69 [0.62;0.77] |
| *Bourgogne-Franche-Comté* | 2.81 [2.58;3.06] | 2.34 [1.96;2.82] | 1.08 [0.90;1.30] | 0.76 [0.62;0.94] | 0.68 [0.58;0.80] |
| *Bretagne* | 2.81 [2.49;3.17] | 1.36 [1.06;1.76] | 1.35 [1.07;1.73] | 0.82 [0.63;1.06] | 0.63 [0.49;0.81] |
| *Centre-Val de Loire* | 2.81 [2.38;3.32] | 3.02 [2.42;3.81] | 1.85 [1.52;2.26] | 0.85 [0.67;1.08] | 0.69 [0.58;0.82] |
| *Grand Est* | 2.81 [2.69;2.93] | 2.01 [1.76;2.32] | 1.15 [1.00;1.32] | 0.60 [0.51;0.70] | 0.73 [0.67;0.81] |
| *Hauts-de-France* | 2.81 [2.60;3.04] | 2.29 [1.92;2.73] | 1.51 [1.31;1.74] | 0.85 [0.72;1.02] | 0.68 [0.61;0.77] |
| *Île-de-France* | 2.81 [2.71;2.92] | 2.51 [2.18;2.87] | 1.70 [1.50;1.92] | 0.72 [0.63;0.82] | 0.65 [0.60;0.70] |
| *Normandie* | 2.81 [2.44;3.24] | 2.63 [2.10;3.28] | 1.38 [1.11;1.70] | 0.63 [0.49;0.81] | 0.67 [0.54;0.83] |
| *Nouvelle-Aquitaine* | 2.81 [2.46;3.21] | 3.01 [2.44;3.74] | 1.17 [0.94;1.43] | 0.73 [0.58;0.94] | 0.63 [0.51;0.77] |
| *Occitanie* | 2.81 [2.53;3.13] | 2.43 [2.00;2.95] | 1.34 [1.11;1.61] | 0.55 [0.42;0.71] | 0.65 [0.54;0.79] |
| *Pays de la Loire* | 2.81 [2.40;3.30] | 2.71 [2.10;3.45] | 1.81 [1.48;2.22] | 0.66 [0.51;0.84] | 0.67 [0.55;0.82] |
| *Provence-Alpes-Côte d’Azur* | 2.81 [2.59;3.05] | 2.31 [1.94;2.76] | 1.32 [1.13;1.55] | 0.89 [0.75;1.08] | 0.71 [0.63;0.81] |
| *France* | 2.81 [2.58;3.07] | 2.42 [2.01;2.90] | 1.46 [1.24;1.72] | 0.71 [0.59;0.87] | 0.67 [0.58;0.78] |

**Figure 4:**
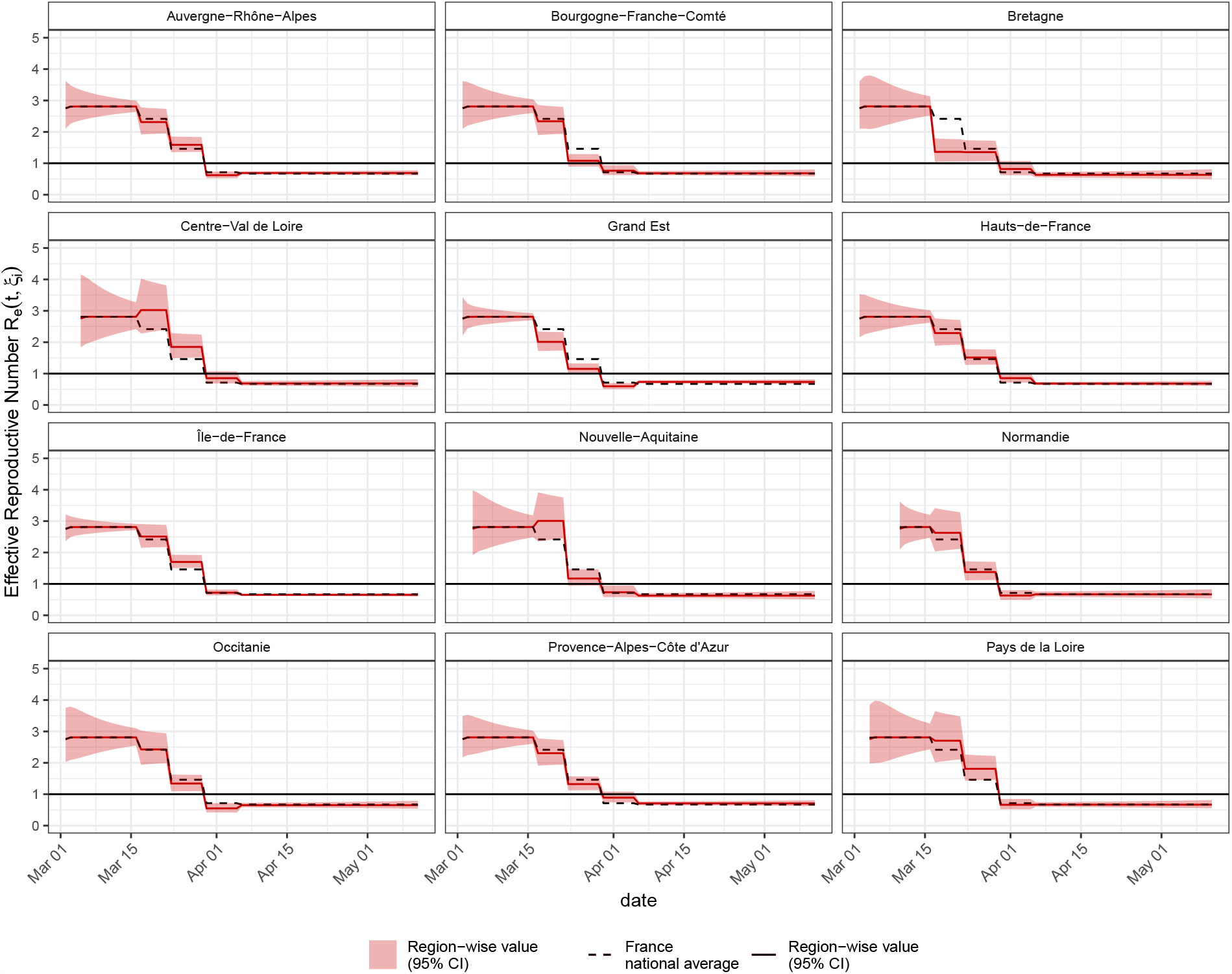
Region specific *R*_*e*_(*t, ξ*_*i*_) compared to the national average. Lockdown started on March 17^th^.

#### Attack rates under lockdown

The map in Figure 5 presents the proportion of infected individuals in the population on 2020-05-11, i.e. instantaneous attack rates. We show that the national French attack rate on May 15^th^ 2020 is 4.9% [3.5%;6.5%]. Table 5 present the attacks rates and their confidence intervals before and after lockdown.

**Figure 5:**
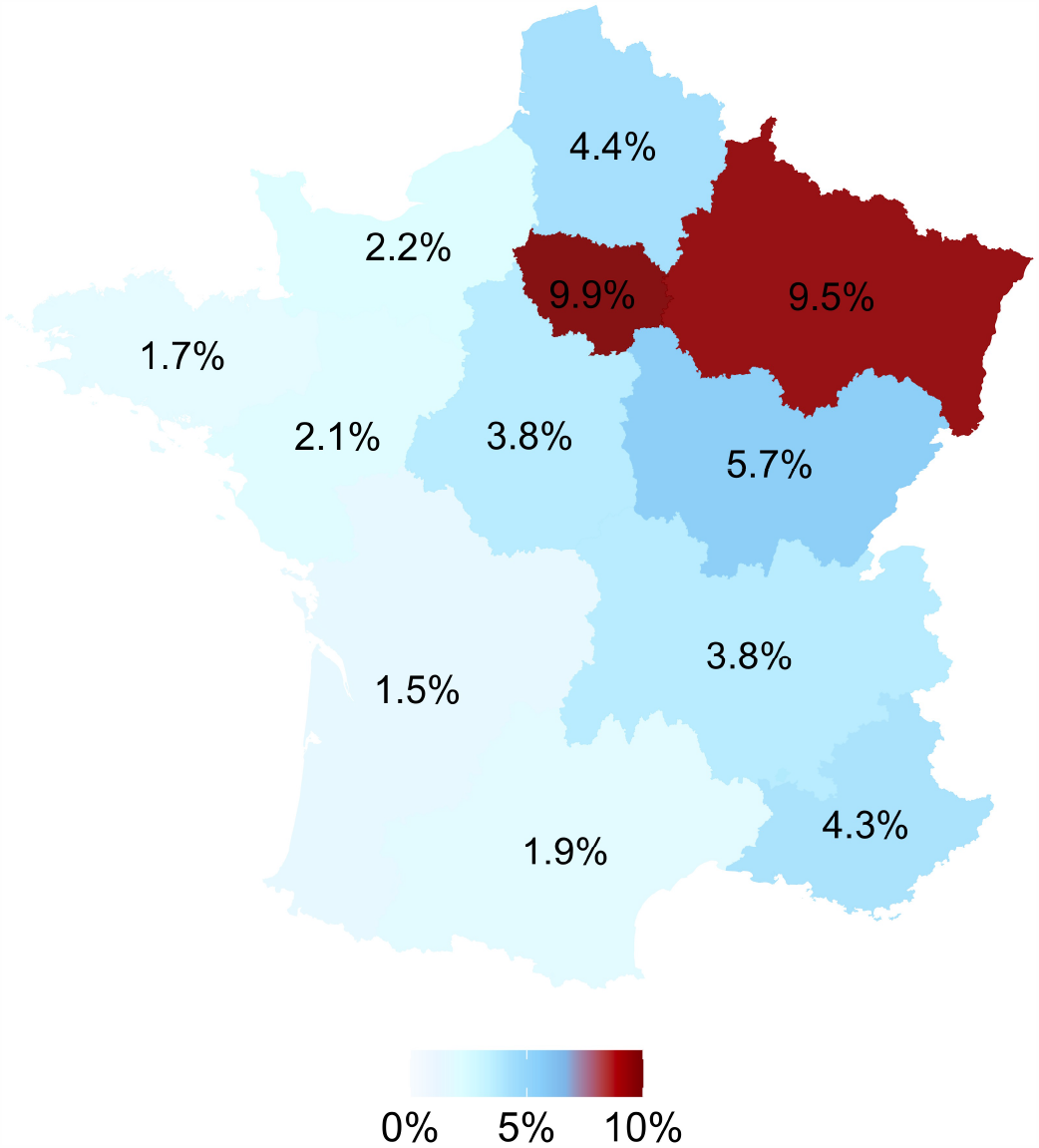
Model estimation for the proportion of Immunized individual in the population (deaths not taken into account), also referred as attack rate, on May 11^th^, 2020.

**Table 5:** Model predictions for the proportion of Infected and Immunized in the population (deaths not taken into account), before and after lockdown, and national weighted averages.

| Region | Attack rates: Infected proportion of the population in % computed as $(E+I+A+H+R)/N$ | |
| --- | --- | --- |
|  | 2020-03-17 | 2020-05-11 |
| *Auvergne-Rhône-Alpes* | 0.45 [0.33;0.55] | 3.80 [2.62; 4.80] |
| *Bourgogne-Franche-Comté* | 0.87 [0.60;1.12] | 5.73 [3.77; 7.53] |
| *Bretagne* | 0.35 [0.14;0.65] | 1.69 [0.72; 3.30] |
| *Centre-Val de Loire* | 0.23 [0.01;0.40] | 3.80 [0.26; 6.53] |
| *Grand Est* | 1.79 [1.61;1.93] | 9.46 [8.05;10.69] |
| *Hauts-de-France* | 0.49 [0.34;0.62] | 4.43 [2.95; 5.96] |
| *Île-de-France* | 1.05 [0.98;1.13] | 9.86 [8.13;12.15] |
| *Nouvelle-Aquitaine* | 0.15 [0.07;0.26] | 1.48 [0.71; 2.55] |
| *Normandie* | 0.23 [0.17;0.32] | 2.23 [1.52; 3.19] |
| *Occitanie* | 0.25 [0.09;0.41] | 1.85 [0.77; 3.10] |
| *Pays de la Loire* | 0.17 [0.03;0.35] | 2.08 [0.40; 4.56] |
| *Provence-Alpes-Côte d’Azur* | 0.51 [0.35;0.67] | 4.29 [2.94; 5.67] |
| *France* | 0.62 [0.48;0.75] | 4.93 [3.51; 6.52] |

#### Optimal lockdown duration

Assuming that the lockdown effect remains constant and equal to *β*_*w*4*i*_ after May 11^th^ 2020, maintaining the lockdown during 180 [150; 215] additional days (i.e. until November 7^th^ 2020) would have been necessary to ensure complete extinction of the epidemics in France, such as compartment E, A and I are empty (i.e. *E* < 1, *A* < 1, and *I* < 1).

## 4 Discussion

In this work, we provide estimations of the key parameters of the dynamics of the COVID-19 epidemic in French regions as well as forecasts according to NPIs especially regarding the proportion of infected when lifting the lockdown policy.

The point estimates of the basic reproductive ratios for French regions fluctuated between 2.4 and 3.4 before lockdown took effect, but according to the uncertainty around these estimates they are not substantially different from one region to another. Therefore, observed differences in the number of cases were due to the epidemic starting first in *Grand Est* and *Île-de-France* regions. These estimates were close to those reported before isolation using other models (Alizon et al., 2020; Flaxman et al., 2020). The model provided estimates of the impact of the lockdown on the effective reproductive ratio with a substantial reduction of *R*_*e*_ after the lockdown. In addition, the model provides estimates of the size of the population of people who have been or are infected as of May 11^th^, 2020. Our estimates for this proportion of subjects ranges from 1.48% to 9.46% of the population in the different regions, thus excluding any herd immunity (control of the epidemic by having a large proportion of people already infected and therefore not susceptible), as similarly reported by Di Domenico et al. (2020) and Salje et al. (2020). With our estimates of basic reproduction ratio and model assumptions, the epidemic would become extinct by herd immunity with a proportion of 89.5% (95% CI [88.0%; 90.7%]) of infected people.

Interpretation of our results is conditional on the mechanistic model illustrated in Figure 1, and careful attention must be given to the parameters set from the scientific literature and detailed in Table 2, as updated estimates are published every day. First and foremost, our model takes only two kinds of infectious cases into account: confirmed cases *I*, and unascertained cases Our observation model takes *I* as the number of infectious cases confirmed by a positive PCR SARS-Cov-2 test. Thus, *A* can be interpreted as unconfirmed symptomatic cases that can be diagnosed by a GP visit (possibly through remote teleconsultation). This is a very simple representation of the COVID-19 infection, which can have various degrees of severity (e.g. asymptomatic, mild, severe) that could be themselves modeled into different compartments. However, very little data is currently available to gather sufficient information to be able to distinguish between those infectious states. Second, our model does not have a compartment for COVID-19 patients in ICU. Compared to Wang et al. (2020), our model does not feature an inflow of susceptibles *n* (and matching outflow) but population movement across regions are limited during the isolation period. Deaths were also not distinguished from recoveries in the *R* compartment, but over the observation period this did not impact the main estimates. Third, our model does not take into account the age-structure of the population on the contrary to the recently posted report Salje et al. (2020) using French data. Interestingly, although the models were different and the data not fully identical, our results were comparable. Actually, our approach captures a part of the unexplained variability between regions through the random effects. This variability might be explained at least partly through the difference in age-structure and probability of hospitalization according to the age, as well as population density and transportation habits.

Based on a model referred in the epidemics literature as a type of cross-coupled metapopulation model (Chowell et al., 2016), our approach additionally incorporate a statistical model on certain parameters and specifically solve the inverse-problem in which we infer those parameters from the available data. Hence, the more data are available the more precise will this parameter estimation be. Thus, we would like to underline the interest of making the data publicly accessible, especially early on during the epidemic growth when information is scarce, as was done by *Santé Publique France* on the data.gouv.fr web portal (https://www.data.gouv.fr/fr/organizations/sante-publique-france/) as well as on the GEODES platform (https://geodes.santepubliquefrance.fr/), hence allowing us to work immediately on this topic. Furthermore, we have made our code fully available on GitHub www.github.com/sistm/SEIRcovid19 for facilitating its dissemination and re-use.

In conclusion, the lockdown has clearly helped controlling the epidemics in France in every region. The number of infected people varies from one region to the other because of the variations in the epidemic start in these regions (both in terms of timing and size). Hence, the predicted proportion of infected people as of May 11 varies, but stays below 10 % everywhere. It is clear from this model, as in other published models (Di Domenico et al., 2020; Flaxman et al., 2020), that a full and instantaneous lockdown lift would lead to a rebound. Additional measures may help in controlling the number of new infections such as strict case isolation, contact tracing (Di Domenico et al., 2020) and certainly a protective vaccine for which the strategy of administration to the population remains to be defined (Amanat and Krammer, 2020; Lurie et al., 2020; Thanh et al., 2020).

## Data Availability

All data and code are available on a github repository

https://www.github.com/sistm/SEIRcovid19

## Acknowledgements

The authors thank Romain Griffier for his time in discussing the aggregated features of COVID-19 patients care at the Bordeaux University Hospital. The authors thank the opencovid-19 initiative for their contribution in opening the data used in this article. BPH thanks Vincent Pey for discussions about the clinical characteristics of the COVID-19 infection. We thank Lixoft SAS for their support. Numerical computations were in part carried out using the PlaFRIM experimental testbed, supported by Inria, CNRS (LABRI and IMB), Université de Bordeaux, Bordeaux INP and Conseil Régional d’Aquitaine (see https://www.plafrim.fr/). This work is supported in part by Inria Mission COVID19, project GESTEPID.

## Availability statement

The data from the SurSaUD^*®*^ database regarding COVID-19 is available from the data.gouv French government platform at https://www.data.gouv.fr/fr/datasets/donnees-des-urgences-hospitalieres-et-de-sos-medecins-relatives-a-lepidemie-de-covid-19 or alternatively on the GEODES platform by *Santé Publique France* (https://geodes.santepubliquefrance.fr). The source code used for this work is available on GitHub at www.github.com/sistm/SEIRcovid19. The Kalman implementation used in Section 2 of Web Appendix 1 is available upon request.

## Author contributions

MP, LW, RT and BPH designed the study. MP and BPH analyzed the data. MP, DD and BPH implemented the software code. QC performed identifiability and asymptotic analysis of the model. AC and PM performed the Kalman-based estimation and implemented the corresponding software. MP, LW, AC, PM, RT and BPH interpreted the results and wrote the manuscript.

